# Reduced Spinal Cord Gray Matter in Patients with Fibromyalgia Using Opioids Long-term

**DOI:** 10.1101/2023.05.02.23289401

**Authors:** Anne K. Baker, Su Hyoun Park, Kenneth A. Weber, Katherine T. Martucci

## Abstract

**Objective:** Chronic pain involves alterations in brain gray matter volume (GMV). Moreover, opioid medications are known to reduce GMV in numerous brain regions involved in pain processing. However, no research has evaluated (1) chronic pain-related GMV alterations in the spinal cord or (2) the effect of opioids on spinal cord GMV. Accordingly, this study evaluated spinal cord GMV in health controls and patients with fibromyalgia who were using and not using opioids long-term.

**Methods:** We analyzed average C5 - C7 GMV of the spinal cord dorsal and ventral horns in separate female cohorts of healthy controls (HC, n = 30), fibromyalgia patients not using opioids (FMN, n = 31), and fibromyalgia patients using opioids long-term (FMO, n = 27). To assess the effect of group on average dorsal and ventral horn GMV, we conducted a one-way multivariate analysis of covariance.

**Results:** After controlling for age, we observed a significant effect of group on ventral horn GMV (*p* = 0.03, η^2^ = 0.09), and on dorsal horn GMV (*p* = 0.05, η^2^ = 0.08). Tukey’s posthoc comparisons showed that, compared to HC participants, FMOs had significantly lower ventral (*p* = 0.01) and dorsal (*p* = 0.02) GMVs. Among FMOs only, ventral horn GMV was significantly positively associated with pain severity and interference, and both dorsal and ventral GMVs were significantly positively associated with cold pain tolerance.

**Conclusion:** Long-term opioid use may impact sensory processing in fibromyalgia via gray matter changes within the cervical spinal cord.

## Introduction

A wide array of chronic pain conditions, including fibromyalgia (FM), involve structural differences in brain gray matter density and volume.^1–7^ The mechanistic underpinnings of these differences remain unclear.^8^ However, structural brain abnormalities yield insight into chronic-pain related adaptations within the central nervous system (CNS). No prior chronic pain research has similarly evaluated gray matter differences in the spinal cord. Within the spinal cord, there are numerous opportunities for modulation of both ascending and descending nociceptive signaling; accordingly, spinal cord structural abnormalities may further represent chronic pain-related alterations within the CNS. Alongside structural brain differences, analysis of structural spinal cord differences may provide lower-order diagnostic and prognostic biomarkers of chronic pain and thereby bolster our understanding of the CNS states that uniquely underlie various chronic pain conditions, including FM.

Reduced gray matter volume (GMV) in the cervical spinal cord has been associated with disability and disease progression in multiple sclerosis,^9,10^ and with clinical disability in amyotrophic lateral sclerosis.^11^ Moreover, clinical outcomes following injury to the spinal cord have been associated with dorsal and ventral horn gray matter atrophy. Specifically, sensory disturbances were associated with dorsal horn atrophy and motor impairments were associated with ventral horn atrophy.^12,13^ Thus, disease processes that include sensory processing disruptions appear to involve gray matter alterations in the dorsal and ventral horns of the cervical spine. Dorsal and ventral horn gray matter alterations have not been examined in FM. However, disrupted sensory processing also appears to involve functional alterations in the dorsal and ventral horns, which have indeed been demonstrated in FM.^14^

Functional magnetic resonance imaging (fMRI) in patients with FM has revealed unbalanced cervical spinal cord activity. Compared to healthy controls, patients with FM exhibit (1) greater ventral and lesser dorsal resting-state activity,^14^ and (2) lesser dorsal horn activity during experimental heat pain.^15^ Together, these findings suggest functional imbalances between the ventral and dorsal horns of the cervical spinal cord may be implicated in the development and maintenance of FM. In particular, central sensitization—a persistent state of heightened CNS activity^16,17^—may in part be a function of increased ventral and decreased dorsal horn activity. Interestingly, prescription opioid medications may partially normalize these imbalances. FM patients who take opioids exhibit ventral and dorsal horn activity that is more similar to healthy controls, while FM patients who don’t take opioids exhibit more pronounced regional differences.^18^

To summarize, *structural* spinal cord changes have been observed in diseases that involve sensory processing disruptions, but not yet in FM. However, *functional* spinal cord imbalances have been observed in FM, and these imbalances may be reduced by prescription opioid use in patients with FM. In light of these considerations, we posited that FM may involve dorsal and ventral horn GMV alterations. Furthermore, given that opioid medications reduce gray matter volumes in brain regions involved in pain processing,^19–21^ we hypothesized that opioids may likewise impact spinal cord dorsal and ventral horn GMV. Due to the dearth of research examining spinal cord GMV differences in FM (or in any other chronic pain condition), we did not posit a directional hypothesis; instead, we hypothesized that we would observe significant group differences between FM patients taking opioids, FM patients not taking opioids, and pain-free healthy controls.

## Patients and Methods

### Participants

Data were obtained from female healthy controls and females with fibromyalgia (N = 96) at Duke University. Six subjects were excluded due to missing scan data and 2 subjects were excluded for poor data quality. Thus, 88 participants were included in the final analysis: 30 healthy control participants (HC), 27 patients with fibromyalgia who were taking opioids (FMO), and 31 patients with fibromyalgia who were not taking opioids (FMN). HC participants had no history of chronic pain and no history of substance abuse, including no extended (>30 days) or recent (within the past 90 days) opioid use. Participants with FM were eligible if they met modified American College of Rheumatology 2016 criteria for fibromyalgia (Wolfe et al., 2016). These criteria consisted of (1) a widespread pain index (WPI) of ≥ 7 plus a symptom severity (SS) score ≥ 5, or WPI score 3-6 plus SS score ≥ 9, (2) comparable symptoms present for at least 3 months, and (3) no diagnosis that would otherwise explain the pain. Additionally, participants were required to have pain in all 4 body quadrants, an average 0-10 verbal pain scale rating ≥ 2, and no uncontrolled psychiatric disorders. FMN participants were free of any substance use (confirmed by urine drug screen immediately prior to scanning) and endorsed no lifetime history of extended (> 30 days) or recent (within the past 90 days) opioid use. FMO participants reported current, regular use of opioids for at least the past 90 days. Opioid medication use and non-opioid medication use (e.g., anti-depressants, acetaminophen) are reported in **Table 1**.

**Table 1.** Non-opioid Pain and Mood-Altering Medication Use.

| **Medication** | **FMO (n)** | **FMN (n)** | **HC (n)** |
| --- | --- | --- | --- |
| Tapentadol | 2 | 0 |  |
| Tramadol | 11 | 1 |  |
| NSAIDs | 9 | 8 |  |
| Acetaminophen | 10 | 5 |  |
| Other Pain Meds (e.g., lidocaine) | 1 | 1 | 1 |
| SNRIs | 9 | 7 |  |
| SSRIs | 4 | 7 |  |
| Tricyclic Antidepressants | 2 | 1 |  |
| Other Anxiolytics | 4 | 2 |  |
| Triptans | 1 | 3 |  |
| Other SRIs | 5 | 2 |  |
| NDRIs | 3 | 5 |  |
| SR Antagonists | 3 | 1 |  |
| Antiepileptics | 8 | 2 |  |
| Benzodiazepines & Benzodiazepine-like | 5 | 8 | 1 |
| Muscle Relaxants | 12 | 8 |  |
| GABA Analogs | 10 | 5 |  |
| Oral Contraceptives | 3 | 10 | 5 |

Participants self-reported regular use of any pain and/or mood-altering medications. Qualitative responses were transformed into binary yes (1) – no (0) responses and summed for all yes answers in each substance category. FMO = opioid-using fibromyalgia, FMN = non-opioid-using fibromyalgia, HC = healthy control. NSAID = non-steroidal anti-inflammatory, SNRI = selective norepinephrine reuptake inhibitor, SSRI = selective serotonin reuptake inhibitor, SRI = serotonin reuptake inhibitor, NDRI = norepinephrine-dopamine reuptake inhibitor, SR = serotonin receptor, GABA = $\gamma$ -Aminobutyric acid.

### Study Procedures

All participants provided written informed consent prior to study procedures, and all study protocols were approved by Duke University’s Institutional Review Board.

#### MRI Acquisition and Preprocessing

All study procedures were conducted in the Brain Imaging and Analysis Center at Duke University. Participants completed demographic and clinical questionnaires, as well as screening for MRI contraindications, prior to scanning. Imaging data were collected with a GE Signa Premier 3.0 Tesla scanner equipped with a 21-channel head and neck neurovascular array coil. Two structural scans were acquired: a T2-weighted sequence extending from the top of the cerebellum to the bottom of the T1 vertebra (single slab 3D fast spin echo, repetition time [TR] = 2500 ms, echo time [TE] = 85 ms, echo train length 70, field of view [FOV] = 240 × 240 mm^2^, matrix size = 256 × 256, slice thickness = 1.4 mm, effective resolution = 1.4 × 0.94 × 0.94 mm^3^, interpolated resolution = 0.7 × 0.47 × 0.47 mm^3^, number of averages = 2), and a 2D T2*-weighted axial multi-echo gradient-echo (MERGE) sequence acquired perpendicular to the spinal cord centered at the C6 vertebra (flip angle 20°, TE = 5.4 ms, TR = 525 ms, number of echoes = 3, 32 oblique slices, FOV = 180 × 144 mm^2^, matrix size = 320 × 192, in-plane resolution = 0.35 × 0.35 mm^2^, slice thickness = 3mm, 0.5 mm spacing, number of averages = 2).^22^

Image preprocessing was performed using Spinal Cord Toolbox (SCT) 5.0.1.^23^ The spinal cord segmentation was obtained using the sct_deepseg_sc command, then manually checked for quality control and updated as necessary. A invertebral disc levels mask was then created by manually identifying the posterior aspect of each intervertebral disc (C2/C3, C3/C4, C4/C5, C5/C6, C6/C7, C7/T1, T1/T2) on the T2-weighted images, which was subsequently used to register the T2-weighted images to the PAM50 T2-weighted template. The sct_deepseg_gm command was then used to segment the spinal cord gray matter, which was then registered to the PAM50 gray matter template using T2-to-template warps to initialize the T2* registration. The PAM50 atlas was then warped to the T2*-weighted images and the gray matter volumes for ventral and dorsal horns (left and right combined) were extracted from the top of the C5 vertebra through the bottom of the C7 vertebra ^23^ **(Figure 1)**. Total horn volumes were then calculated as left and right sums for each horn, and mean ventral and dorsal volumes per axial slice were computed by dividing horn size (voxels) by the number of slices.

**Figure 1.**
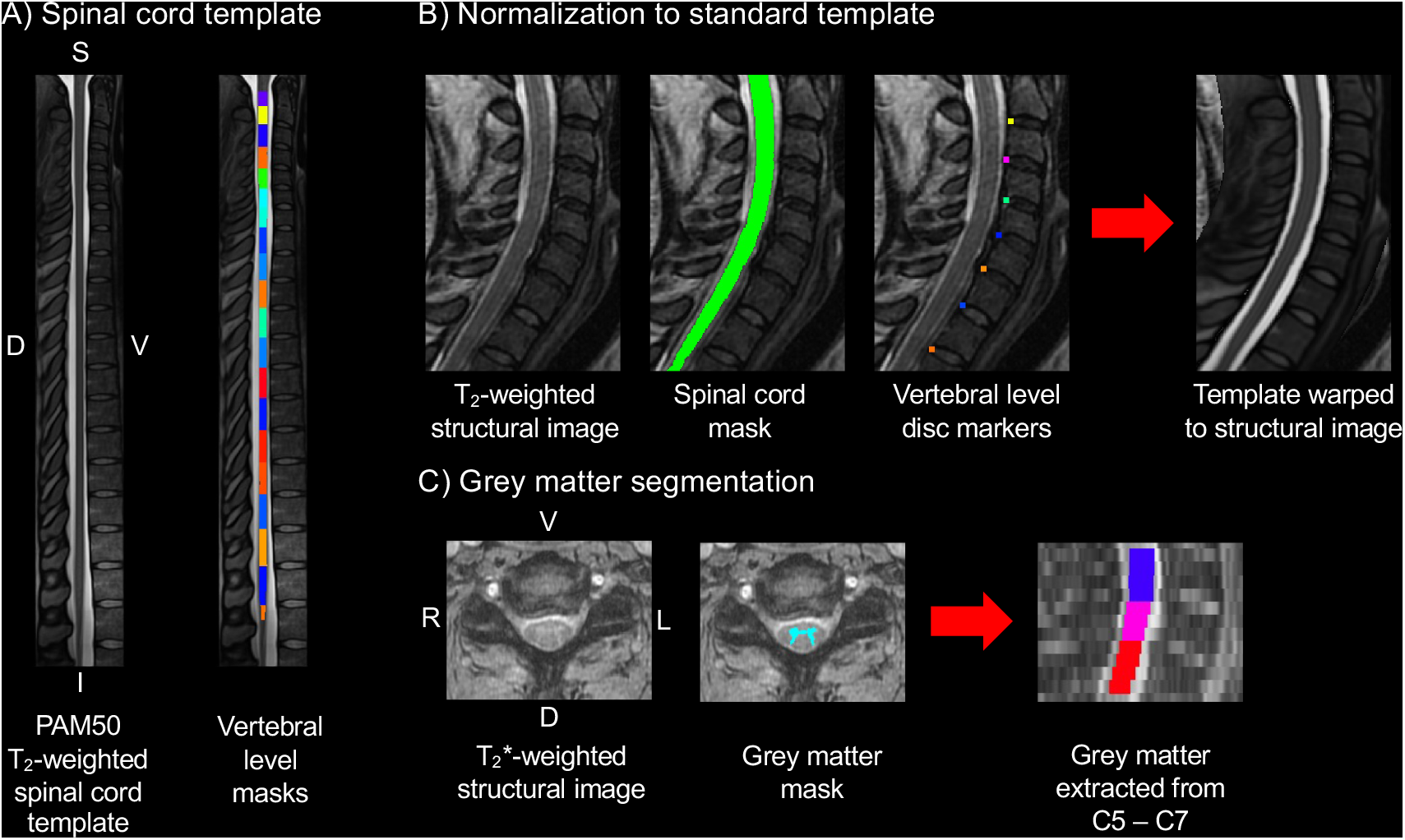
Spinal cord preprocessing was performed using the Spinal Cord Toolbox. Images from a healthy control were used to create the graphics for this figure. **Panel A** shows the PAM50 T_2_-weighted template and the vertebral level masks. **Panel B** shows the template to T_2_-weighted structural image registration process. **Panel C** shows the process of segmenting the spinal cord gray matter. Gray matter was extracted from the top of the C5 vertebra to the bottom of the C7 vertebra, and total horn volumes were computed as left and right sums for each horn. D = Dorsal, V = Ventral, S = Superior, I = Inferior, L = Left, R = Right.

#### Quantitative Sensory Testing

Quantitative Sensory Testing (QST) was performed following the MRI scan, including the Trapezius Pressure Pain Threshold (PPT) Test and the Cold Pressor Test (CPT)—both of which are validated measures of central sensitization.^24,25^ The PPT is a test of mechanical pain sensitivity evoked by gradually increasing applied pressure. A hand-held pressure algometer was used for the PPT Test (FPK10, Wagner Instruments, Greenwich, CT, USA). For the test, pressure was applied to the trapezius muscle at the point halfway between the neck and the lateral aspect of the shoulder. Participants verbally indicated the point at which the pressure first became painful. An initial practice test was first conducted (throw-away value). Then, pressure was applied three times (or until repeated tests’ responses were within 0.2 kgf) per side to the left and right trapezius muscles. Average pressure (in kg/cm^2^) across the three stimulations were computed as the final metrics representing bilateral pressure pain thresholds. The CPT is a test of cold pain tolerance. Participants submerged their left hand into a container of cold water (approximately 5°C) for up to 2 minutes. Total time (seconds) immersed in cold water served as the final metric representing cold pain tolerance.

### Statistical Analysis

All statistical analyses were conducted using IBM SPSS 26 (IBM Corporation, Armonk, NY, USA). Between-group differences between FMO, FMN and HC participants were evaluated with an independent samples t-test. To assess the effect of group on average dorsal and ventral horn gray matter volumes, we conducted a one-way multivariate analysis of covariance (MANCOVA). Dorsal and ventral horn gray matter volumes were entered as dependent variables, group was entered as a fixed factor, and age was entered as a covariate of no interest. Prior to analysis, Shapiro-Wilk tests revealed that dorsal horn volumes among HC participants were non-normally distributed. Removal of one outlier allowed for HC participants to meet this assumption, and we proceeded with the planned analysis. Post-hoc pairwise comparisons were corrected for multiple comparsions with Fisher’s least significant difference (LSD) procedure. Pearson’s bivariate associations—stratified by group—were conducted to examine relationships between dorsal horn and ventral horn gray matter volumes and (1) QST measures (PPT, CPT), (2) pain questionnaires (pain severity and interference from the Brief Pain Inventory^26^), (3) morphine dosage (MME), and (4) pain duration (years). Bivariate correlations were Bonferroni corrected for multiple comparisons.

## Results

### General Characteristics

Baseline characteristics of FM patients and corresponding HC participants are shown in **Table 2**. Group differences in age were evaluated using one-way analysis of variance. We observed significant differences in age (F_(2,52)_ = 4.52, *p* = 0.02), and Tukey’s post-hoc comparisons showed age was significantly higher among opioid-using FM participants (FMO) relative to non-opioid using FM patients (FMN; *p* = 0.01). Significant group differences were also observed in CPT submersion times (F_(2,81)_ = 3.51, *p* = 0.04) and in average bilateral pain pressure thresholds (F_(2,81)_ = 22.98, *p* < 0.001). Tukey’s post-hoc comparisons showed that HC participants exhibited (1) significantly greater CPT submersion time compared to FMN participants (*p* = 0.03), and (2) significantly higher pain pressure thresholds than both FM cohorts (*p* < 0.001 for both). Finally, we also observed a significant difference in pain duration between the FM cohorts (t_(55)_ = -3.53, *p* < 0.001).

**Table 2.** Group Differences in Baseline Characteristics.

|  | **FMO**<br>**Mean (SD)** | **FMN**<br>**Mean (SD)** | **HC**<br>**Mean (SD)** | **t/F** | **p** |
| --- | --- | --- | --- | --- | --- |
| Age | 49.35<br>(9.53) | 36.41<br>(11.82) | 45.61<br>(12.55) | 9.09 | **0.0003\*\*** |
| Pain Duration (yrs) | 11.98<br>(6.97) | 6.41<br>(4.88) |  | -3.53 | **<0.001\*\*\*** |
| Pain Severity | 5.75<br>(1.25) | 5.44<br>(1.13) |  | -0.93 | 0.18 |
| Pain Interference | 6.49<br>(2.34) | 6.93<br>(2.00) |  | 0.73 | 0.24 |
| CPT Submersion<br>Time (secs) | 52.87<br>(43.39) | 33.31<br>(29.90) | 59.84<br>(43.88) | 3.51 | **0.03\*** |
| PPT | 1.48<br>(0.65) | 1.61<br>(0.49) | 3.36<br>(1.82) | 22.98 | **<0.001\*\*\*** |

Between-group differences were evaluated with an independent samples t-test and one-way analysis of variance. FMO = opioid-using fibromyalgia, FMN = non-opioid-using fibromyalgia, HC = healthy control. CPT = cold pressor test, PPT = pain pressure threshold.

### Cervical Spinal Cord (C5-C7) Gray Matter Volume Analyses

After controlling for age, we observed a significant effect of group on ventral horn gray matter volume (F_(2,78)_ = 3.89, *p* = 0.03, η^2^ = 0.09), and on dorsal horn gray matter volume (F_(2,78)_ = 3.17, *p* = 0.05, η^2^ = 0.08). We examined post-hoc pairwise comparisons with Tukey’s multiple comparison test. Pairwise comparisons showed that, compared to healthy controls, FMO participants had significantly lower ventral (*p* = 0.01, 95% CI [-588.81, -97.14]) and dorsal (*p* = 0.02, 95% CI [55.94, 527.27]) horn gray matter volumes. No other pairwise comparsions were significant. Mean volumes for each group are reported in **Table 3**.

**Table 3.**
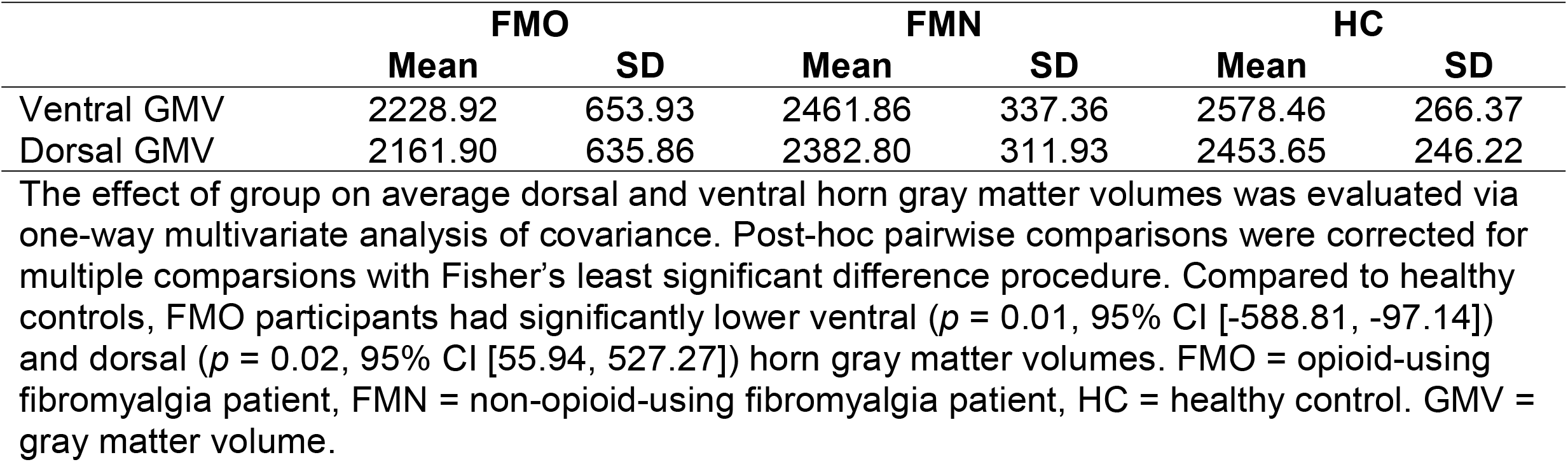
Mean C5-C7 Dorsal and Ventral Horn Volumes Stratified by Group.

### Bivariate Associations

To gain further insight into the extent to which dorsal horn gray matter volume may be related to pain sensitivity, we examined bivariate associations—stratified by group—between average dorsal and ventral gray matter volumes, QST measures, and clinical measures of pain. Significant correlations were found only among the FMO cohort, as follows. Ventral horn gray matter volume was significantly positively associated with pain severity (*p* = 0.005) and pain interference (*p* = 0.006) **(Figure 2)**. Both ventral and dorsal horn gray matter volumes were significantly positively associated with CPT submersion time (*p* = 0.002, *p* = 0.006, respectively) **(Figure 3)**. Correlations between dorsal horn gray matter volume, pain severity and pain interference trended toward significance, but did not meet the significance threshold after correcting for multiple comparisons. Among FMN participants, correlations between dorsal and ventral horn gray matter volumes and pain duration also trended toward significance, but again did not meet the significance threshold after correcting for multiple comparisons. All other bivariate correlations were non-significant **(Table 4)**.

**Table 4.**
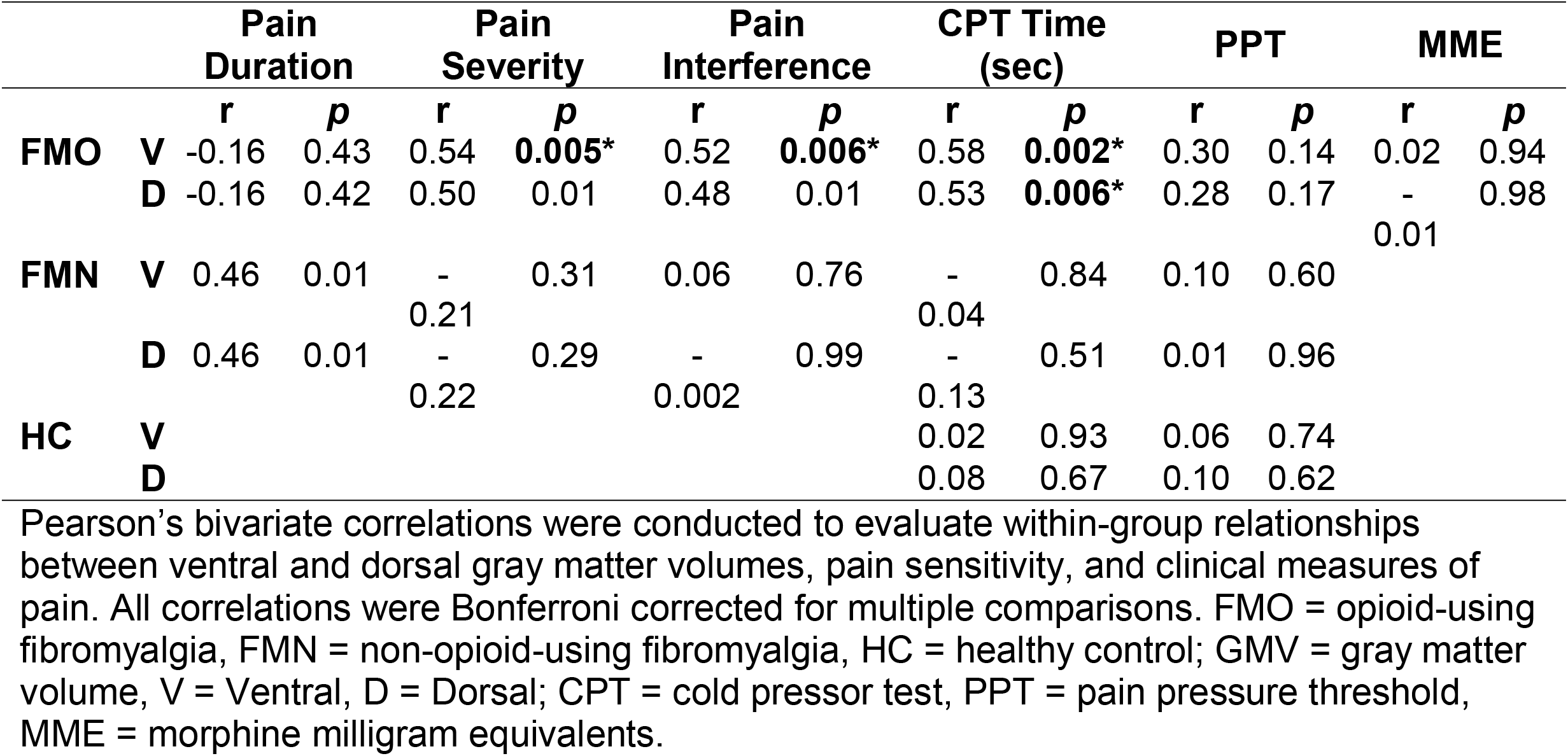
Pearson’s Bivariate Correlations Between Average C5-C7 Dorsal and Ventral Gray Matter Volumes, Pain Measures, and Average Opioid Medication Dose.

**Figure 2.**
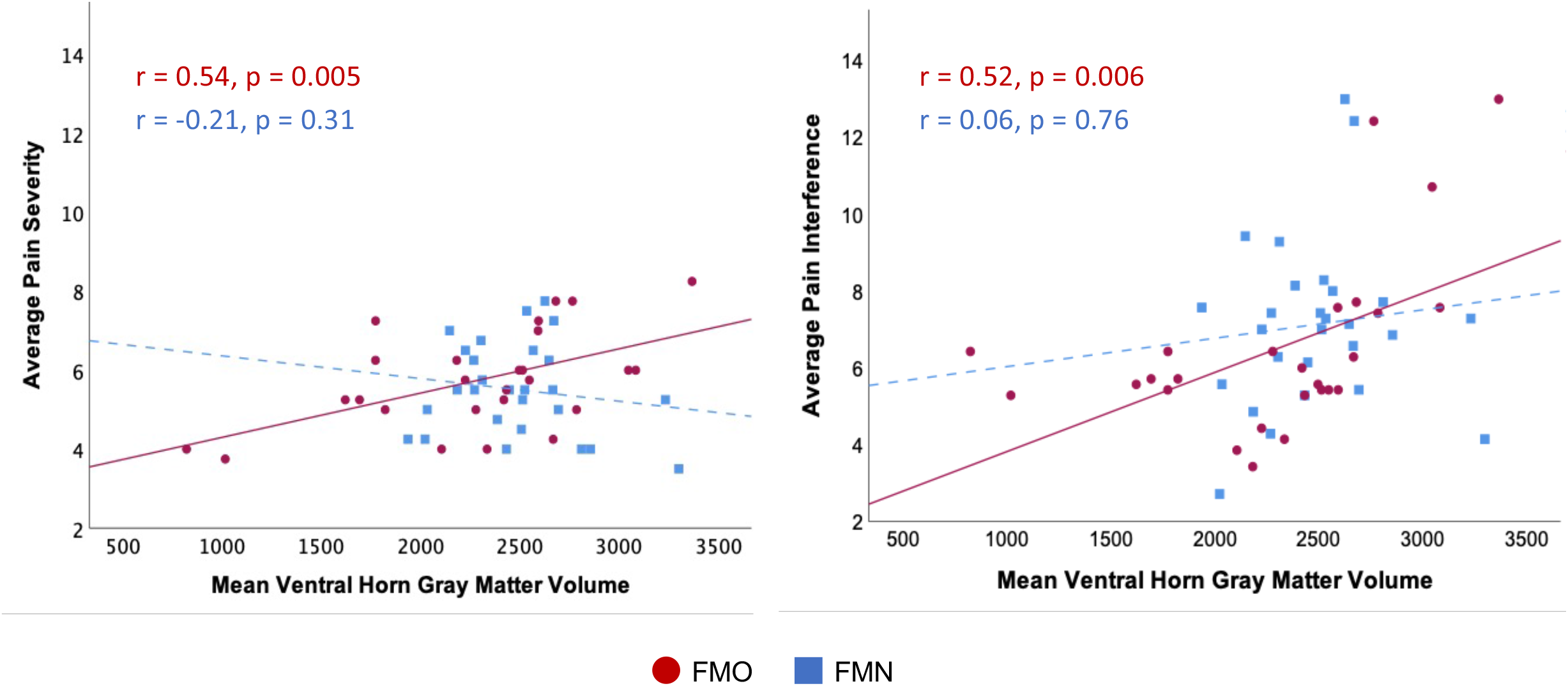
Group-stratified bivariate associations between mean C5 – C7 ventral horn gray matter volume, average pain severity, and average pain interference. Bivariate correlations were Bonferroni corrected for multiple comparisons. FMO = opioid-using fibromyalgia patients; FMN = non-opioid-using fibromyalgia patients.

**Figure 3.**
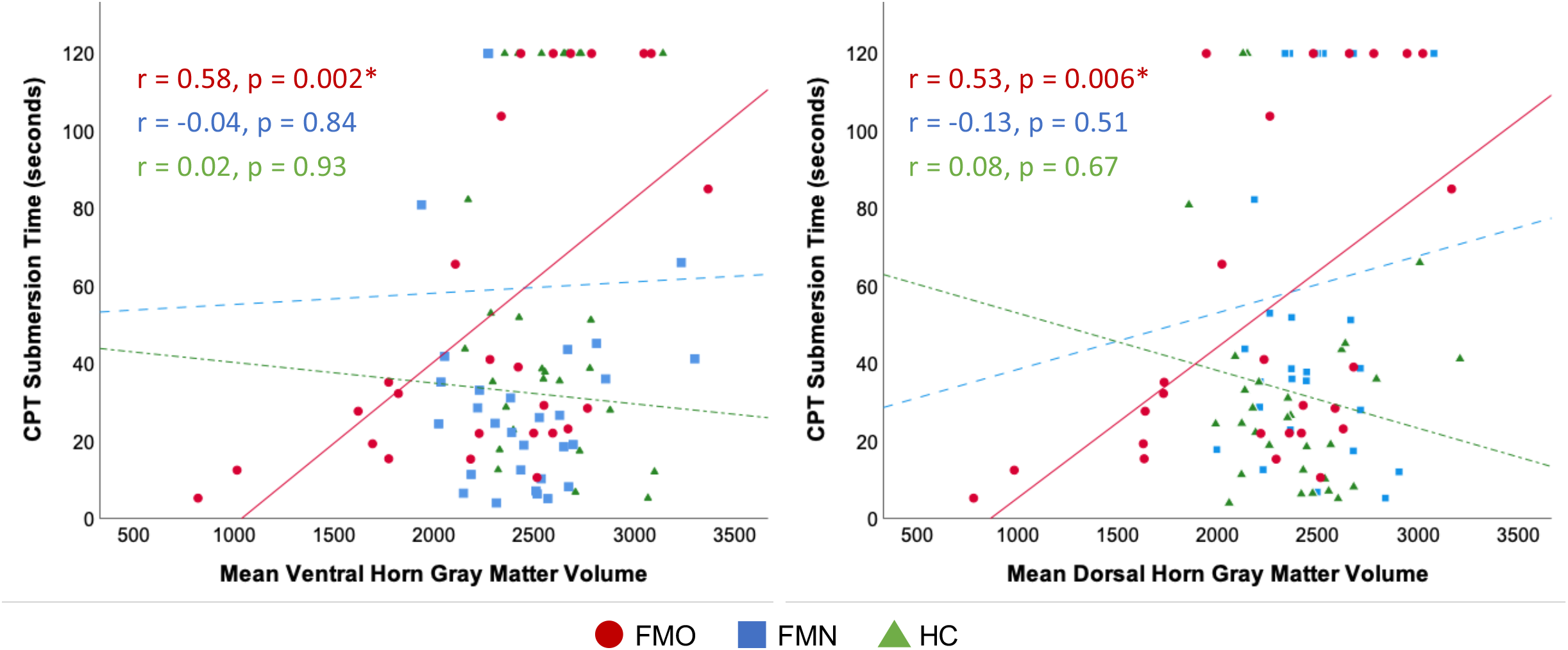
Group-stratified bivariate associations between mean C5 – C7 ventral and dorsal horn gray matter volumes and cold pressor test (CPT) submersion time in seconds. Bivariate correlations were Bonferroni corrected for multiple comparisons. FMO = opioid-using fibromyalgia patients, FMN = non-opioid-using fibromyalgia patients, HC = healthy controls.

## Discussion

The present study provides the first examination of cervical spinal cord gray matter in FM. We present novel evidence that long-term opioid use appears to significantly impact gray matter volume in both the ventral and dorsal horns of the cervical spinal cord. Our results build upon prior functional spinal cord research that reveals ventral-dorsal imbalances in FM,^14^ and potential opioid-related normalization of those imbalances.^18^ Together, these studies highlight the importance of evaluating (1) chronic pain-related structural changes throughout the entire central nervous system, and (2) the extent to which long-term opioid use may be associated with spinal cord gray matter alterations in chronic pain.

Overall, relative to healthy controls, mean dorsal and ventral GMVs were lower among both patient cohorts. This observation in the cervical spinal cord is consistent with previous reports of brain gray matter decreases associated with FM.^6,7,27–29^ However, statistically significant differences in dorsal and ventral GMVs were only observed between opioid-using patients and healthy controls. This finding in the spinal cord is consistent with prior brain research that demonstrates long-term opioid use in the context of chronic pain is associated with decreased regional brain gray matter volumes.^19–21^

There are a few possible explanations for the present findings. Multimodal imaging of the brain has previously been used to test whether regional brain GMV decreases associated with FM are undergirded by compromised neuronal integrity, as measured by GABA_A_ receptor concentration.^7^ Importantly, regional brain GMV decreases did not appear to be explained by GABA_A_ receptor concentration, but instead by decreased tissue water content—theoretically as a function of altered cerebral blood flow, which has been previously observed in FM.^7,30^ It is plausible that, similar to prior brain research, decreased water content may likewise account for the reduced cervical spinal cord ventral and dorsal horn GMVs observed among opioid-using FM patients in the present study.

Meanwhile, it is important to note that this prior multimodal brain research did not specifically examine relationships between opioid use and reduced regional GMVs in FM. If the results of the present study were to truly parallel prior explorations of FM-related reductions in brain GMV, it seems that we would have observed significantly lower GMVs in both fibromyalgia cohorts. Certainly, the fact that dorsal and ventral GMV was significantly lower only among opioid-using FM patients requires further consideration. This finding may be at least partially attributable to significant differences in pain duration, as opioid-using FM patients had signifncatly longer average pain duration. It is possible that some temporal threshold exists at which pain-related GMV changes in the spinal cord begin to become detectable via MRI, and that significant GMV differences observed among the FMO cohort are merely representative of longer pain duration. Decreased tissue water content could also be attributable to dehydration, secondary to long-term opioid use.^31^

Alternatively, lower ventral and dorsal horn GMV uniquely observed in the FMO cohort could be attributable to opioid-related neuronal changes—specifically, lower concentrations of GABA. Opioids inhibit GABA-mediated synaptic transmission.^32,33^ While this is partially what contributes to the analgesic effects of opioids, it may also be the case that opioids disrupt a critical excitatatory and inhibitory balance in the spinal cord. This hypothesis may provide a framework in which to understand our observation that significantly lower ventral and dorsal horn GMV among the FMO cohort was also significantly positively correlated with pain severity and interference.

However, this hypothesis does not aid in understanding our finding that CPT submersion times were signficantly positively associated with GMVs only among the FMO cohort, nor why CPT submersion time was higher among FMOs than among FMNs. While the positive association between GMVs and pain intensity and interference would suggest opioids may have a deleterious effect in FM, the positive correlation between GMVs and CPT submersion time would suggest the opposite. One possible explanation for this is that CPT submersion time is heavily influenced by psychological factors (e.g., anxiety, expectancy, positive affect).^34–36^ It may be that opioids are acting on affective factors that improve tolerance to pain, while simultaneously acting on sensory processing mechanisms that paradoxically increase pain. Overall, our bivariate results underscore the value of assessing within-group analyses alongside between-group analyses, and future research should aim to continue identifying not only between-group differences in spinal cord gray matter volumes, but also the extent to which those differences are clinically meaningful in different cohorts.

A few limitations should be noted in consideration of these results. First, this study is cross-sectional and thus does not afford causal inferences regarding the impact of opioids on spinal cord gray matter volumes over time. Future longitudinal studies initiated at commencement of opioid therapy will be essential to more accurately and explicitly capture the discrete effects of opioids on ventral and dorsal GMVs. Additionally, this study was conducted in an all-female sample. Pain and opioid responses vary across the menstrual cycle, and sex hormones may significantly modify responses to opioids.^37^ As such, future studies that capture the effects of menstural-cycle related variability are critical to validate and extend the results of the present study. Furthermore, replication studies in cohorts of both males and females are likewise necessary to assess sex-specific changes in chronic pain, which have been previously described in the brain.^38^ In a similar vein, future studies of cervical spinal cord gray matter alterations in other chronic pain conditions will also be critical to assess the generalizability of the present results. Finally, despite being adequately powered to control for age, we were unable to additionally control for the statistically significant difference in pain duration observed between FM cohorts. Accordingly, further research is needed to explore the impact of pain duration on spinal cord GMV alterations in FM. Likewise, the statistically significant difference in age between FM cohorts also requires future research to explore, rather than control for, the impact of aging on spinal cord GMV.

Overall, the present study contributes novel evidence of opioid-related reductions in cervical spinal cord dorsal and ventral horn GMV in patients with FM on long-term opioid therapy. Our findings hold valuable implications for future mechanistic studies of FM—and of chronic pain in general—particularly regarding the extent to which opioids may adaptively or maladaptively alter spinal cord structural integrity in chronic pain. We posit that opioids may alter ventral and dorsal GMV and concomitant pain through inhibition of GABA-mediated synaptic transmission. Future longitudinal and multimodal imaging studies will be essential to further evaluate this hypothesis.

## Data Availability

All data produced in the present study are available upon reasonable request to the authors.

## References

1. Bagarinao E, Johnson KA, Martucci KT, Ichesco E, Farmer MA, Labus J, Ness TJ, Harris R, Deutsch G, Apkarian VA, Mayer EA, Clauw DJ, Mackey S. Preliminary structural MRI based brain classification of chronic pelvic pain: A MAPP network study. Pain. 2014 Dec;155(12):2502–2509. PMCID: PMC4504202

2. Kairys AE, Schmidt-Wilcke T, Puiu T, Ichesco E, Labus JS, Martucci K, Farmer MA, Ness TJ, Deutsch G, Mayer EA, Mackey S, Apkarian AV, Maravilla K, Clauw DJ, Harris RE. Increased brain gray matter in the primary somatosensory cortex is associated with increased pain and mood disturbance in patients with interstitial cystitis/painful bladder syndrome. J Urol. Ovid Technologies (Wolters Kluwer Health); 2015 Jan;193(1):131–137.

3. Apkarian AV, Sosa Y, Sonty S, Levy RM, Harden RN, Parrish TB, Gitelman DR. Chronic back pain is associated with decreased prefrontal and thalamic gray matter density. J Neurosci. 2004 Nov 17;24(46):10410–10415. PMCID: PMC6730296

4. Barad MJ, Ueno T, Younger J, Chatterjee N, Mackey S. Complex regional pain syndrome is associated with structural abnormalities in pain-related regions of the human brain. J Pain. Elsevier; 2014 Feb;15(2):197–203. PMCID: PMC4784981

5. Shi H, Yuan C, Dai Z, Ma H, Sheng L. Gray matter abnormalities associated with fibromyalgia: A meta-analysis of voxel-based morphometric studies. Semin Arthritis Rheum. 2016 Dec;46(3):330–337. PMID: 27989500

6. McCrae CS, O’Shea AM, Boissoneault J, Vatthauer KE, Robinson ME, Staud R, Perlstein WM, Craggs JG. Fibromyalgia patients have reduced hippocampal volume compared with healthy controls. J Pain Res. 2015 Jan 30;8:47–52. PMCID: PMC4321661

7. Pomares FB, Funck T, Feier NA, Roy S, Daigle-Martel A, Ceko M, Narayanan S, Araujo D, Thiel A, Stikov N, Fitzcharles MA, Schweinhardt P. Histological Underpinnings of Grey Matter Changes in Fibromyalgia Investigated Using Multimodal Brain Imaging. J Neurosci. 2017 Feb 1;37(5):1090–1101. PMCID: PMC6596849

8. Martucci KT, Mackey SC. Neuroimaging of Pain: Human Evidence and Clinical Relevance of Central Nervous System Processes and Modulation. Anesthesiology. 2018 Jun;128(6):1241–1254. PMCID: PMC5953782

9. Schlaeger R, Papinutto N, Panara V, Bevan C, Lobach IV, Bucci M, Caverzasi E, Gelfand JM, Green AJ, Jordan KM, Stern WA, von Büdingen HC, Waubant E, Zhu AH, Goodin DS, Cree BAC, Hauser SL, Henry RG. Spinal cord gray matter atrophy correlates with multiple sclerosis disability. Ann Neurol. 2014 Oct;76(4):568–580. PMCID: PMC5316412

10. Agosta F, Pagani E, Caputo D, Filippi M. Associations between cervical cord gray matter damage and disability in patients with multiple sclerosis. Arch Neurol. jamanetwork.com; 2007 Sep;64(9):1302–1305. PMID: 17846269

11. Paquin MÊ, El Mendili MM, Gros C, Dupont SM, Cohen-Adad J, Pradat PF. Spinal Cord Gray Matter Atrophy in Amyotrophic Lateral Sclerosis. AJNR Am J Neuroradiol. 2018 Jan;39(1):184–192. PMCID: PMC7410702

12. Huber E, David G, Thompson AJ, Weiskopf N, Mohammadi S, Freund P. Dorsal and ventral horn atrophy is associated with clinical outcome after spinal cord injury. Neurology. 2018 Apr 24;90(17):e1510–e1522. PMCID: PMC5921039

13. Smith ZA, Weber KA 2nd, Paliwal M, Hopkins BS, Barry AJ, Cantrell D, Ganju A, Koski TR, Parrish TB, Dhaher Y. Magnetic Resonance Imaging Atlas-Based Volumetric Mapping of the Cervical Cord Gray Matter in Cervical Canal Stenosis. World Neurosurg. 2020 Feb;134:e497–e504. PMCID: PMC7024652

14. Martucci KT, Weber KA, Mackey SC. Altered Cervical Spinal Cord RestingCState Activity in Fibromyalgia. Arthritis & Rheumatology [Internet]. Wiley Online Library; 2019; Available from: https://onlinelibrary.wiley.com/doi/abs/10.1002/art.40746

15. Bosma RL, Mojarad EA, Leung L, Pukall C, Staud R, Stroman PW. FMRI of spinal and supra-spinal correlates of temporal pain summation in fibromyalgia patients. Hum Brain Mapp. 2016 Apr;37(4):1349–1360. PMCID: PMC4783193

16. Woolf CJ. Central sensitization: implications for the diagnosis and treatment of pain. Pain. 2011 Mar;152(3 Suppl):S2–S15. PMCID: PMC3268359

17. Latremoliere A, Woolf CJ. Central sensitization: a generator of pain hypersensitivity by central neural plasticity. J Pain. 2009 Sep;10(9):895–926. PMCID: PMC2750819

18. Martucci KT, Weber KA 2nd, Mackey SC. Spinal Cord Resting State Activity in Individuals With Fibromyalgia Who Take Opioids. Front Neurol. 2021 Aug 4;12:694271. PMCID: PMC8371264

19. Upadhyay J, Maleki N, Potter J, Elman I, Rudrauf D, Knudsen J, Wallin D, Pendse G, McDonald L, Griffin M, Anderson J, Nutile L, Renshaw P, Weiss R, Becerra L, Borsook D. Alterations in brain structure and functional connectivity in prescription opioid-dependent patients. Brain. 2010 Jul;133(Pt 7):2098–2114. PMCID: PMC2912691

20. Younger JW, Chu LF, D’Arcy NT, Trott KE, Jastrzab LE, Mackey SC. Prescription opioid analgesics rapidly change the human brain. Pain. 2011 Aug;152(8):1803–1810. PMCID: PMC3138838

21. Lin JC, Chu LF, Stringer EA, Baker KS, Sayyid ZN, Sun J, Campbell KA, Younger JW. One Month of Oral Morphine Decreases Gray Matter Volume in the Right Amygdala of Individuals with Low Back Pain: Confirmation of Previously Reported Magnetic Resonance Imaging Results. Pain Med. 2016 Aug;17(8):1497–1504. PMCID: PMC4921346

22. Cohen-Adad J, Alonso-Ortiz E, Abramovic M, Arneitz C, Atcheson N, Barlow L, Barry RL, Barth M, Battiston M, Büchel C, Budde M, Callot V, Combes AJE, De Leener B, Descoteaux M, de Sousa PL, Dostál M, Doyon J, Dvorak A, Eippert F, Epperson KR, Epperson KS, Freund P, Finsterbusch J, Foias A, Fratini M, Fukunaga I, Wheeler-Kingshott CAMG, Germani G, Gilbert G, Giove F, Gros C, Grussu F, Hagiwara A, Henry PG, Horák T, Hori M, Joers J, Kamiya K, Karbasforoushan H, Keřkovský M, Khatibi A, Kim JW, Kinany N, Kitzler H, Kolind S, Kong Y, Kudlička P, Kuntke P, Kurniawan ND, Kusmia S, Labounek R, Laganà MM, Laule C, Law CS, Lenglet C, Leutritz T, Liu Y, Llufriu S, Mackey S, Martinez-Heras E, Mattera L, Nestrasil I, O’Grady KP, Papinutto N, Papp D, Pareto D, Parrish TB, Pichiecchio A, Prados F, Rovira À, Ruitenberg MJ, Samson RS, Savini G, Seif M, Seifert AC, Smith AK, Smith SA, Smith ZA, Solana E, Suzuki Y, Tackley G, Tinnermann A, Valošek J, Van De Ville D, Yiannakas MC, Weber KA 2nd, Weiskopf N, Wise RG, Wyss PO, Xu J. Generic acquisition protocol for quantitative MRI of the spinal cord. Nat Protoc. 2021 Oct;16(10):4611–4632. PMID: 34400839

23. De Leener B, Lévy S, Dupont SM, Fonov VS, Stikov N, Louis Collins D, Callot V, Cohen-Adad J. SCT: Spinal Cord Toolbox, an open-source software for processing spinal cord MRI data. Neuroimage. 2017 Jan 15;145(Pt A):24–43. PMID: 27720818

24. Hazra S, Venkataraman S, Handa G, Yadav SL, Wadhwa S, Singh U, Kochhar KP, Deepak KK, Sarkar K. A Cross-Sectional Study on Central Sensitization and Autonomic Changes in Fibromyalgia. Front Neurosci. 2020 Aug 4;14:788. PMCID: PMC7417433

25. Finan PH, Buenaver LF, Bounds SC, Hussain S, Park RJ, Haque UJ, Campbell CM, Haythornthwaite JA, Edwards RR, Smith MT. Discordance between pain and radiographic severity in knee osteoarthritis: findings from quantitative sensory testing of central sensitization. Arthritis Rheum. 2013 Feb;65(2):363–372. PMCID: PMC3863776

26. Cleeland CS, Ryan KM. Pain assessment: global use of the Brief Pain Inventory. Ann Acad Med Singapore. 1994 Mar;23(2):129–138. PMID: 8080219

27. Ceko M, Bushnell MC, Fitzcharles MA, Schweinhardt P. Fibromyalgia interacts with age to change the brain. Neuroimage Clin. 2013 Sep 6;3:249–260. PMCID: PMC3814958

28. Kuchinad A, Schweinhardt P, Seminowicz DA, Wood PB, Chizh BA, Bushnell MC. Accelerated brain gray matter loss in fibromyalgia patients: premature aging of the brain? J Neurosci. 2007 Apr 11;27(15):4004–4007. PMCID: PMC6672521

29. Robinson ME, Craggs JG, Price DD, Perlstein WM, Staud R. Gray matter volumes of pain-related brain areas are decreased in fibromyalgia syndrome. J Pain. 2011 Apr;12(4):436–443. PMCID: PMC3070837

30. Williams DA, Gracely RH. Biology and therapy of fibromyalgia. Functional magnetic resonance imaging findings in fibromyalgia. Arthritis Res Ther. 2006;8(6):224. PMCID: PMC1794529

31. Mallappallil M, Sabu J, Friedman EA, Salifu M. What Do We Know about Opioids and the Kidney? Int J Mol Sci [Internet]. 2017 Jan 22;18(1). Available from: http://dx.doi.org/10.3390/ijms18010223 PMCID: PMC5297852

32. Vaughan CW, Ingram SL, Connor MA, Christie MJ. How opioids inhibit GABA-mediated neurotransmission. Nature. 1997 Dec 11;390(6660):611–614. PMID: 9403690

33. Cohen GA, Doze VA, Madison DV. Opioid inhibition of GABA release from presynaptic terminals of rat hippocampal interneurons. Neuron. 1992 Aug;9(2):325–335. PMID: 1497896

34. Lee JE, Watson D, Frey Law LA. Lower-order pain-related constructs are more predictive of cold pressor pain ratings than higher-order personality traits. J Pain. 2010 Jul;11(7):681–691. PMCID: PMC2904871

35. Feldner MT, Hekmat H. Perceived control over anxiety-related events as a predictor of pain behaviors in a cold pressor task. J Behav Ther Exp Psychiatry. 2001 Dec;32(4):191–202. PMID: 12102581

36. Hanssen MM, Vancleef LMG, Vlaeyen JWS, Peters ML. More optimism, less pain! The influence of generalized and pain-specific expectations on experienced cold-pressor pain. J Behav Med. 2014 Feb;37(1):47–58. PMID: 23239369

37. Ribeiro-Dasilva MC, Shinal RM, Glover T, Williams RS, Staud R, Riley JL 3rd, Fillingim RB. Evaluation of menstrual cycle effects on morphine and pentazocine analgesia. Pain. 2011 Mar;152(3):614–622. PMCID: PMC3039079

38. Baker AK, Ericksen LC, Koppelmans V, Mickey BJ, Martucci KT, Zubieta JK, Love TM. Altered Reward Processing and Sex Differences in Chronic Pain. Front Neurosci. 2022 Jun 7;16:889849. PMCID: PMC9211769

